# Prevalence of Depression and Associated Socio-economic Outcomes during Violent Conflict: A Matched Analysis for Palestine Using Nationally Representative Survey and Conflict Event Data

**DOI:** 10.1101/2024.02.23.24303259

**Authors:** Piero Ronzani, Wolfgang Stojetz, Nadine Stammel, Maria Boettche, Diego Zardetto, Sarah Fenzl, Maen Salhab, Jessica M. Anderson, Arden Finn, Alia Aghajanian, Tilman Brück

## Abstract

**Background:** Mental health risks are high in conflict settings, but mental health research mostly focuses on non-conflict settings. Survey data from active conflict settings often suffer from low response rates, unrepresentative samples, and a lack of detailed information on the roots and implications of poor mental health. We overcome these challenges by analyzing nationally representative evidence on the prevalence, sources, and socio-economic correlates of depression, a highly disabling and costly public health issue, in an active conflict setting.

**Methods:** We analyze nationally and sub-nationally representative geocoded survey data from the Palestinians’ Psychological Conditions Survey, collected from 5,877 Palestinian individuals in West Bank and Gaza in 2022. We calculate representative depression statistics, disaggregate by sub-areas and across socio-demographic groups, and estimate the associations with geocoded violent conflict event data as well as survey-based trauma exposure across conflict types and socio-economic outcomes.

**Findings:** 58 percent (SE=2·21) of adults in Palestine exhibit depressive symptoms. Prevalence is highest in Gaza (71 percent, SE=2·70), increases with exposure to violent conflict and traumatic events, and is associated with worse socio-economic outcomes. The associated losses for 2022 are equivalent to 732,555 Years Lost in Disability, representing 8·9 percent of Palestine’s GDP.

**Interpretation:** Those exposed to violence and traumatic events are disproportionately affected by depression in conflict settings, which may fuel poverty and instability. Scalable investments in mental health in conflict settings promise to not only support well-being but also strengthen productivity and social cohesion for a given level of violence.

**Funding:** The study received funding by the World Bank’s State and Peace Building Trust Fund.

## Research in context

### Evidence before this study

Mental health risks are particularly high in conflict settings, but mental health research has mostly focused on non-conflict or post-conflict settings.

Existing survey data from active conflict settings suffer from low response rates, unrepresentative and small samples, exclusively self-reported conflict exposure, and a lack of detailed information on the roots and implications of poor mental health.

As a result, estimates of the prevalence of mental health issues are non-representative and imprecise and of limited use for assessing socio-economic correlates of poor mental health in active conflict settings.

### Added value of the study

We analyze nationally representative geocoded survey data from the Palestinians’ Psychological Conditions Survey, collected from a sample of 5,877 Palestinian individuals in Palestine in 2022, spatiotemporally matched with disaggregated conflict event data.

We present evidence on the prevalence, sources, and socio-economic correlates of depression, a highly disabling and costly public health issue, in an active conflict setting.

Specifically, we offer nationally representative depression statistics, disaggregated by sub-areas and across socio-demographic groups, quantify the associations of depression with conflict and trauma exposure, externally reported and self-reported respectively, as well as with socio-economic outcomes, and estimate the associated losses in terms of Years Lived with Disability and Gross Domestic Product.

The high-quality, representative, large sample with fine-grained geocodes allows us to produce estimates that are precise. The findings are of high relevance to other populations in active conflict settings.

### Implications of all the available evidence

The study emphasizes the enormous mental health risks that active conflicts entail, as a result of the various adverse conditions and traumatic experiences in violent conflict situations.

The findings in this study also highlight the importance of studying depression, conflict and socioeconomic outcomes jointly, to better and more comprehensively understand challenges to well-being, productivity and social cohesion in low– and middle-income countries.

Further, the findings emphasize the high potential of mental health interventions in active conflict settings, which promise to not only support well-being but also strengthen productivity and social cohesion, for a given level of violence.

## Introduction

Depression is the single most important contributor to Years Lived with Disability (YLD) and the fourth leading contributor to the global burden of disease.^1^ Depression not only creates symptoms such as an inability to feel pleasure, pessimism, disrupted sleep, and poor nutrition but also has important socio-economic consequences at the individual, household, and societal levels. For example, depression can impair economic decision-making and productivity,^2,3,4,5^ compromise the well-being and prospects of affected persons’ children^6^, and reduce social participation.^7^

As of 2019, an estimated 280 million individuals globally were living with depressive disorders.^1^ Among those, 82 percent were located in low– and middle-income countries (LMICs) with estimates suggesting that an individual in a LMIC is 1·5 to 3 times more prone to depression compared to an individual in a high-income setting.^3,7^ Among LMICS, conflict-affected situations merit particular attention as these are the settings where the world’s poorest are increasingly concentrated; adverse conditions and traumatic experiences conducive to depression are particularly prevalent; and access to treatment is very limited.^8,9,10,11,12,13^

To date, there is limited knowledge about depression in conflict settings (as reviewed in Appendix A1 in more detail). The existing research has four limitations. First, while mental health risks are particularly high in conflict settings, mental health research has disproportionately focused on *non-conflict settings.*^2,6^ Second, while there is suggestive evidence that mental health risks are higher during a conflict,^13^ most investigations of conflict-affected settings are limited to *post-conflict situations.*^14^ Third, while there is growing awareness that conditions of scarcity and adversity can strongly affect decision-making and socio-economic outcomes via psychological channels including depression,^3^ existing behavioral research has focused on poor populations in non-conflict settings. In consequence, insights into how *depression affects socio-economic outcomes in conflict settings* are very limited. Fourth, *data collection* in conflict settings faces important challenges.^15,16^ As a result, mental health studies in active conflict settings draw on small convenience samples, response rates are notoriously moderate compromising data quality, and conflict exposure is self-reported. In consequence, estimates of the prevalence of mental health issues are usually non-representative and imprecise while conflict exposure is measured subjectively and may be distorted for endogenous reasons.^9,11,17^

To overcome these four limitations, we collected and analyzed nationally and sub-nationally representative geocoded survey data in the Palestinians’ Psychological Conditions Survey, including 5,877 Palestinian individuals in 2022, following the acute conflict in Gaza in May 2021. We calculate representative depression statistics for Palestine, disaggregate by sub-areas and across socio-demographic groups, quantify the associations of depression with conflict (using precise geocodes) and trauma exposure as well as socio-economic outcomes and estimate the associated losses in terms of YLD and GDP.

Even before the outbreak of the ongoing Israel-Hamas war triggered by the attacks of October 7, 2023, Palestinians faced enduring social, political and economic challenges. Ongoing tensions have led to intermittent escalations, occasionally culminating in major events, such as nine months before the data collection in May 2021.^19^ However, the current war marks a significant escalation of the level of violence in the region, with the number of airstrikes in Gaza exceeding those recorded in a single month for any country or territory in the Middle East since 2020.^20^ Armed clashes in the West Bank also reached an all-time high in October 2023.^20^ Given the current situation, the estimates presented here represent a lower bound of the mental health and socio-economic situation of Palestinians for the foreseeable future.

## Method

### Procedure and participants

This analysis is based on the Palestinians’ Psychological Conditions Survey (PPCS) data collected by the Palestinian Central Bureau of Statistics (PCBS) with technical support by the authors in face-to-face interviews in March and April 2022, almost one year after the conflict in May 2021. Appendix A2 provides detailed information on the sampling design and estimation methodology. In summary, the selection of PPCS households followed a one-stage cluster sampling design. The sample consists of Palestinian residents of the West Bank and Gaza and is representative at the national, governorate, and rural/urban/camp levels. Despite the challenging setting, 87 percent of the 7,057 sampled households were successfully interviewed. Of those who could not be interviewed, in 84 percent of cases the household was not found or had moved abroad, in 12 percent of cases the household refused to be interviewed again, and the remaining households only partially completed the interview. Out of the 6,140 respondent households, 5,877 adult individuals completed all questionnaire modules. The An-Najah National University Institutional Review Board (IRB) approved the present study protocol.

### Measures

We present a short summary of our key measures below and provide more detail in Appendix A3.

#### Depression

To assess the risk of depression, we included the WHO-5 well-being index in the survey.^21^ The WHO-5 was initially developed by the World Health Organisation (WHO) to measure subjective well-being, but has been used as a screening instrument for depression among several studies in recent years. Its reliability and validity to screen for clinical depression has been demonstrated in studies across different countries.^22,23^ The instrument results in a score ranging from 0 to 100. A score below or equal to 50 denotes a screening diagnosis of depression.

We estimate **Years Lived with Disability** (YLD) instead of Disability-adjusted Life Years (DALY), as depression is not categorized as a primary cause of death according to the GBD.^1^ YLD are calculated by the prevalence of individual sequelae of the disease multiplied by their corresponding disability weights.^1^

#### Sociodemographic variables

We study a range of sociodemographic variables and report depression results disaggregated by area of residence (West Bank/Gaza), sex (male/female), education (elementary/high school/university), and refugee status (yes/no). Detailed sociodemographic statistics are reported in Appendix A4.

#### Conflict exposure

We study two indicators of individual conflict exposure,^15^ which we use to proxy the adverse conditions people experience during an armed conflict: 1) self-reported exposure to traumatic events; and 2) spatial and temporal proximity to conflict events based on the Armed Conflict Location & Event Data Project (see www.acleddata.com). For the latter, we compute households’ physical distance to the location of conflict events as recorded in the ACLED dataset.^1^

#### Economic outcomes and well-being

We study four indicators of economic outcomes and well-being concerning perceived poverty and employment. To assess perceived poverty, we use the self-developed survey question “in general, do you consider your household: wealthy/middle/poor/very poor?”. From the answer to this question, we build a measure of the self-assessed degree of poverty. To assess food security, we employ the Food Consumption Score (FCS), a validated dietary diversity indicator with standardized cut-offs that are used across geographic areas and livelihood groups.^24^ For employment, we look at self-reported employment status (employed/unemployed) and for work intensity, we look at hours worked in the week before the interview among those who reported being currently employed at that time.

#### Prosocial behavior

We elicit prosocial behaviors building on previous work probing engagement in prosocial acts.^25,26^ Based on reported engagement in ten examples of prosocial behaviors in the last six months, we categorized participants in quintiles depending on the number of prosocial behaviors.

## Statistical analysis

We conduct univariate and bivariate descriptive analyses based on the matched survey and conflict event data. To ensure representativeness, weights for households and individuals were calculated using a multi-step process designed to reduce the risk of bias and to improve estimation efficiency (see Appendix A2). The procedure involved the following steps: deriving initial weights based on the PPS sampling algorithm, adjusting for non-response at the household level using a response-propensity modeling approach informed by existing panels, calibrating weights against demographic benchmarks, trimming overly large weights to ensure stability of estimates, and finally calibrating individual weights based on household composition for a comprehensive and integrated weighting approach. We report standard 95-percent confidence intervals.

### Role of the funding source

The funding source of the study had no role in study design, data collection, data analysis, data interpretation, or writing of the manuscript. All authors had full access to all the data in the study and had accepted responsibility for the decision to submit for publication.

## Results

### Prevalence of depression

Applying standard WHO-5 criteria, more than half of the adult population of Palestine (58 percent, SE=2·21) have elevated depressive symptom levels. Figure 1 shows the full distribution of depressive symptom levels, the mean level (dashed blue line), and the standard WHO-5 threshold score of 50 (dashed red line). Note that a *lower* score corresponds to a *higher* burden of depression.

**Fig. 1.**
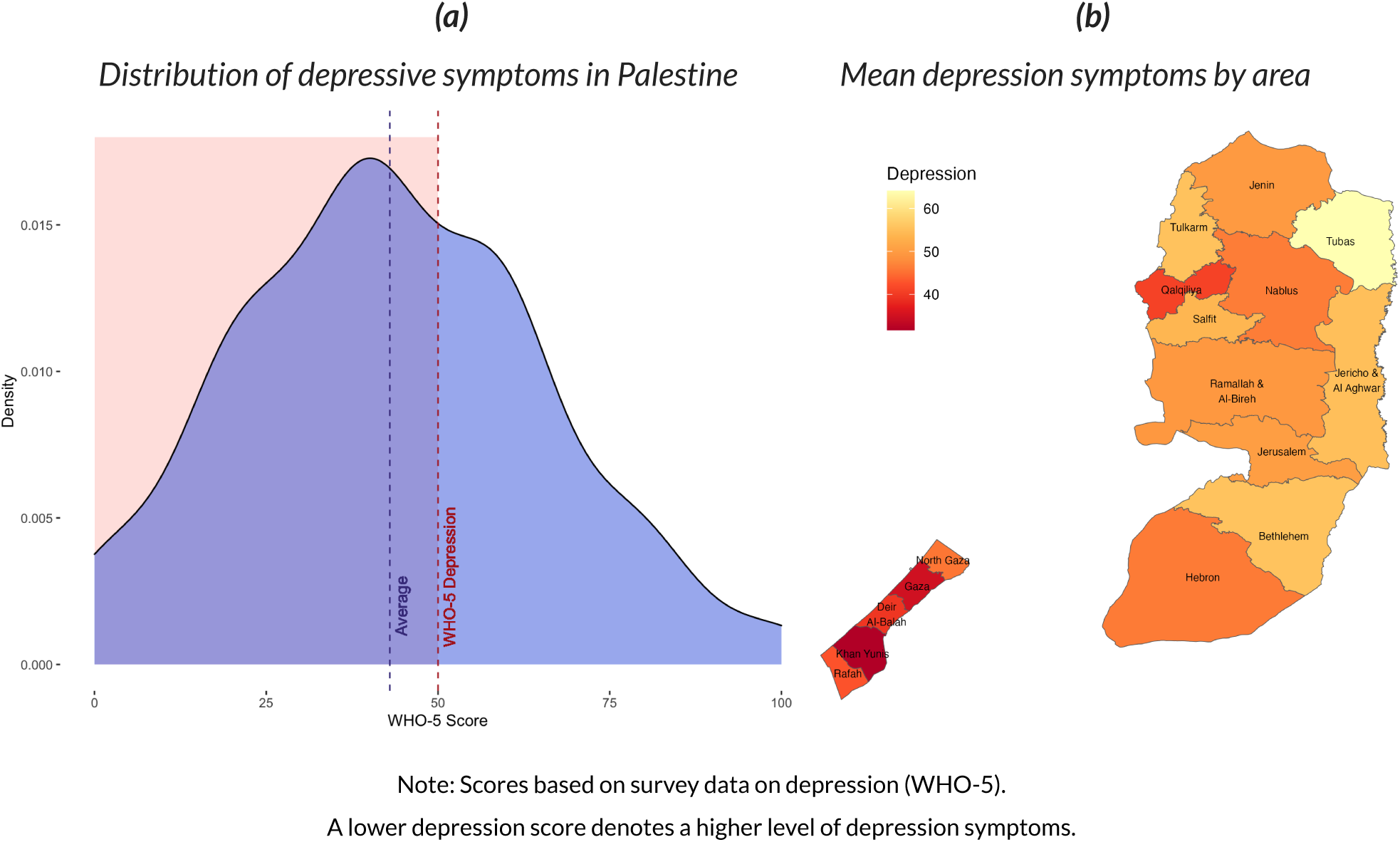
Prevalence of Depression

### Depression by area

As illustrated in Figure 1b, depressive symptom levels vary substantially across different Palestinian governorates. Depressive symptoms are particularly prevalent in Gaza, where 71 percent (SE=2·70) screen positive for depression. In the West Bank, 49 percent (SE=3·05) of the population exhibit depressive symptoms beyond the WHO-5 threshold. The governorates in Gaza register the highest levels of depressive symptoms while the governorate of Qalqiliya exhibits the highest level of depressive symptoms in the West Bank.

### Depression by demographic characteristics

The extent of depression varies significantly across socio-demographic groups defined by individuals’ education level, refugee status, and sex. Figure 2 plots group means separately for Gaza and the West Bank. We find that depression is more common among the less educated (Fig. 2a). For the West Bank, the differences between those who have primary, secondary, and tertiary education (as their highest level of education) are all statistically significant. For Gaza, only the difference between primary and tertiary education is statistically significant. Depression symptoms do not vary significantly between refugees and non-refugees (Fig. 2b). However, depression levels are worse among females than males especially in Gaza (Fig. 2c). Overall, the results suggest that the area of residence (West Bank versus Gaza) is just as important in determining depression levels as other demographic characteristics, emphasizing that in active conflicts local conditions play a key role in shaping mental health outcomes.

**Fig. 2.**
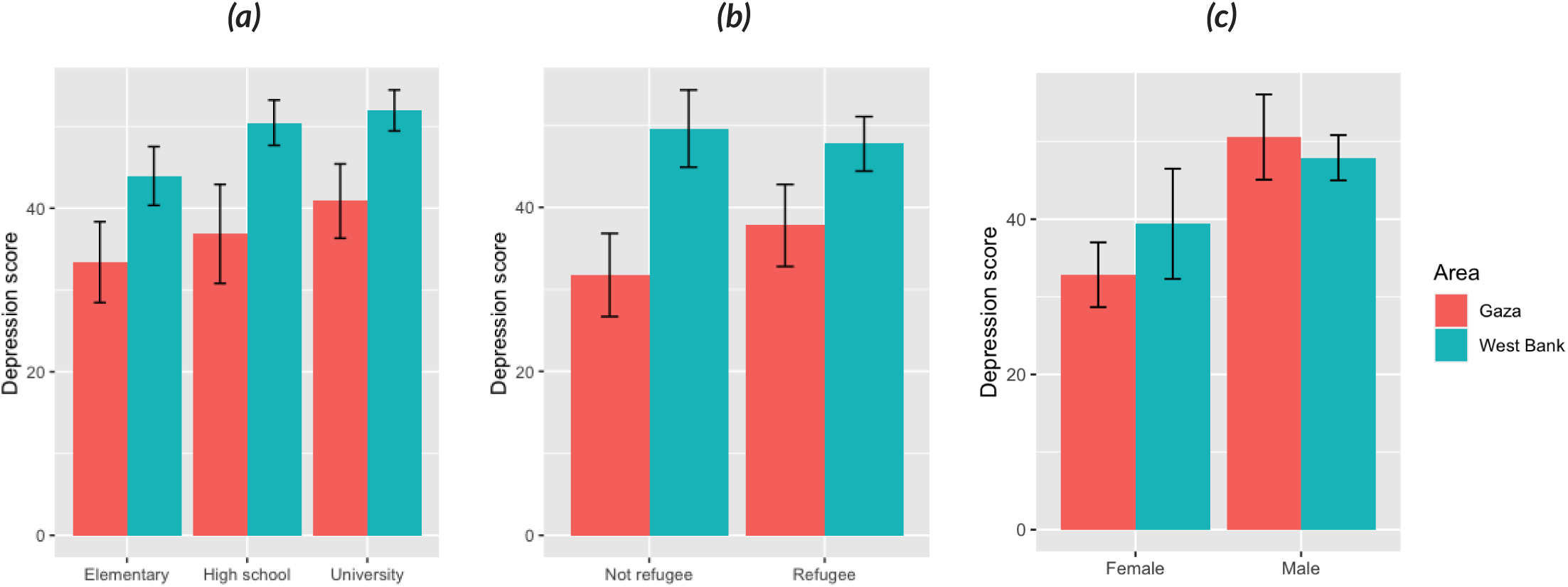
Depression symptoms score (WHO-5) by education (a), refugee status (b), and sex (c), grouped by area of residence

### Depression by exposure to conflict events

Based on conflict event data from the ACLED database, Figures 3a and 3b show the spatial distribution of the intensity of conflict for one year and for five years prior to the survey, respectively. Conflict intensity varies significantly, with the highest levels registered in Gaza City and North Gaza governorates for Gaza and in Jerusalem and Hebron governorates for the West Bank.

**Fig. 3.**
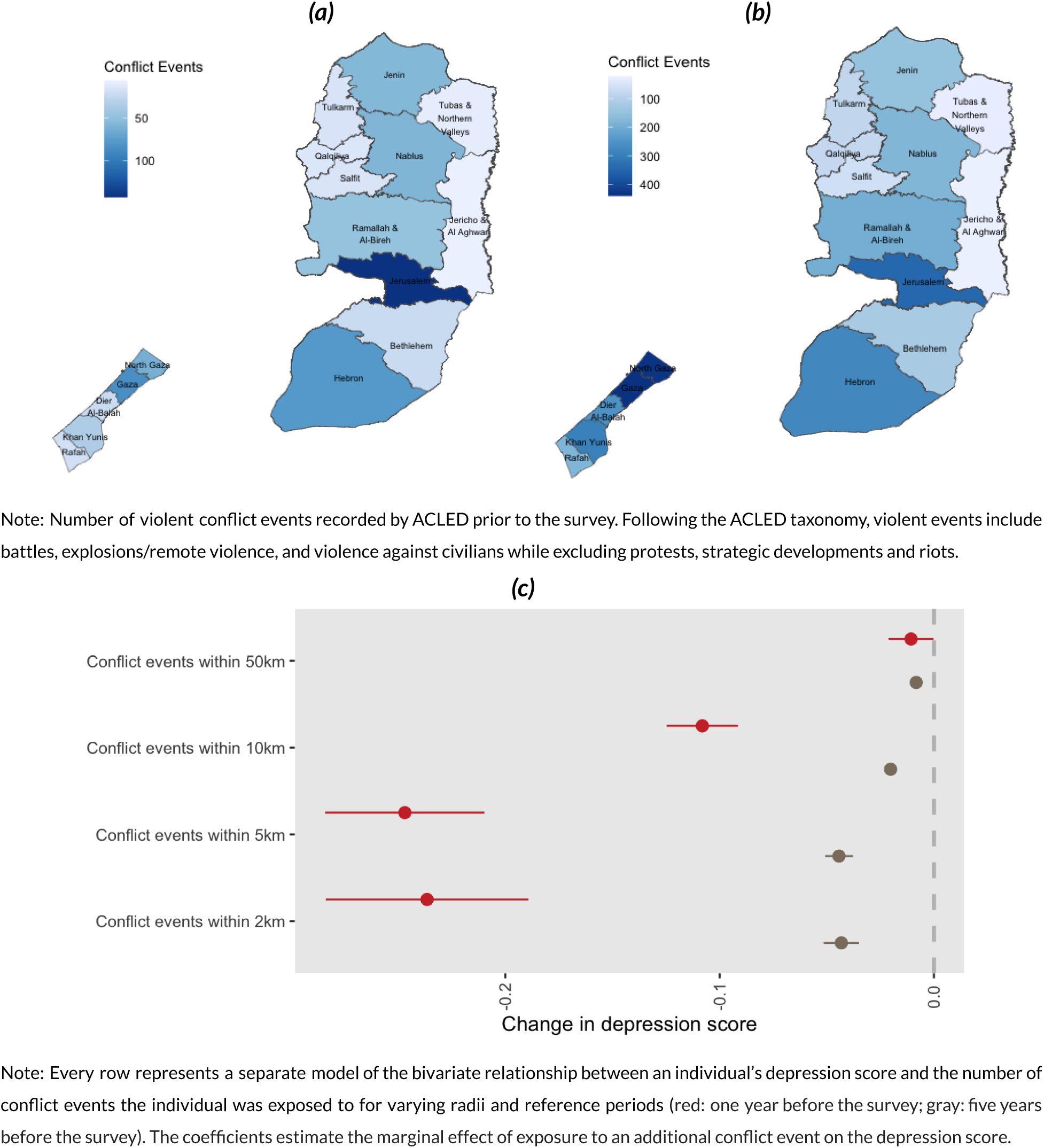
Spatial distribution of conflict events one year (a) and five years (b) before the survey and their relationship with depression (c)

In the year before the survey, individuals were exposed to about 8 events on average in a 2km radius around their home, with a maximum of 65 conflict events (Table A.4 in Appendix A5). During the five years before the survey, individuals were exposed to 45 events on average in a 2km radius around their home, with a maximum of 352 conflict events. These figures increase strongly when we increase the radius around a household’s location. For a 50km radius, individuals were exposed to between 15 and 225 events in the year before the survey (with a mean of 136 events) and between 44 and 1,612 events in the five years before the survey (with a mean of 716 events). These figures emphasize the high levels of conflict intensity many Palestinians are exposed to.

As shown in Figure 3c, we find that the number of conflict events an individual was exposed to is associated significantly with more depressive symptoms. The magnitude of the association is largest for events very close to a household’s location (radii of 2km and 5km). For example, exposure to one additional conflict event in a 2km radius is associated with a ·24 (·04) decrease in the depression score for conflict events in the one year (5 years) before the survey.^2^ That the magnitude of the marginal effect of a single event is significantly smaller for the five-year period is expected because the total number of events individuals were exposed to in the five-year period is much larger than in the one-year period, which is a subset of the five-year period (see Table A.4). Given a mean depression score of around 45, these marginal effects of a single conflict event may seem small at first sight. However, these estimates refer to a single (additional) event while many individuals were exposed to a large number of events even within a 2km radius, as noted above (Table A.4). Therefore, the estimates suggest that exposure to conflict events in recent years may contribute significantly to current depression symptoms.

### Depression by (self-reported) exposure to traumatic events

Even before the outbreak of the conflict in 2023, the prevalence of self-reported trauma exposure in the West Bank and Gaza was very high (Table A.5). For example, 29 percent of Palestinians reported experiencing or witnessed war-related casualties in their lifetime in 2022, and about one in ten Palestinians report having experienced situations involving serious injury or a death threat in their life. Around five percent report having been beaten and six out of every 1,000 Palestinian reported having experienced unwanted sexual contact due to pressure. In Appendix A6, we provide statistics for the full list of all surveyed types of traumatic events in Gaza and the West Bank.

The correlation between the lifetime measure of self-reported trauma exposure and the 5-year measure of event-based conflict exposure consistently increases with the radius considered, from.11 for a 2km radius up to. 18 for a 50km radius (Table A7 in Appendix A7). For the 1-year measure of conflict event exposure there is no such pattern and the correlations are even smaller, as one might expect given the lifetime nature of the trauma exposure measure. These results suggest that the widely-used self-reported trauma exposure measures and event-based conflict exposure measures capture distinct challenges and are thus complements, rather than substitutes, as indicators of conflict exposure, which is an important methodological finding.

Turning to depression, we document a significantly lower WHO-5 score, indicating significantly more depressive symptoms, among those who reported having experienced any traumatic events compared to those who did not (Figure 4). Among those reporting no exposure to traumatic events, the average WHO-5 score is slightly below the cut-off point for depression (M=49·5, SE=2·84).

**Fig. 4.**
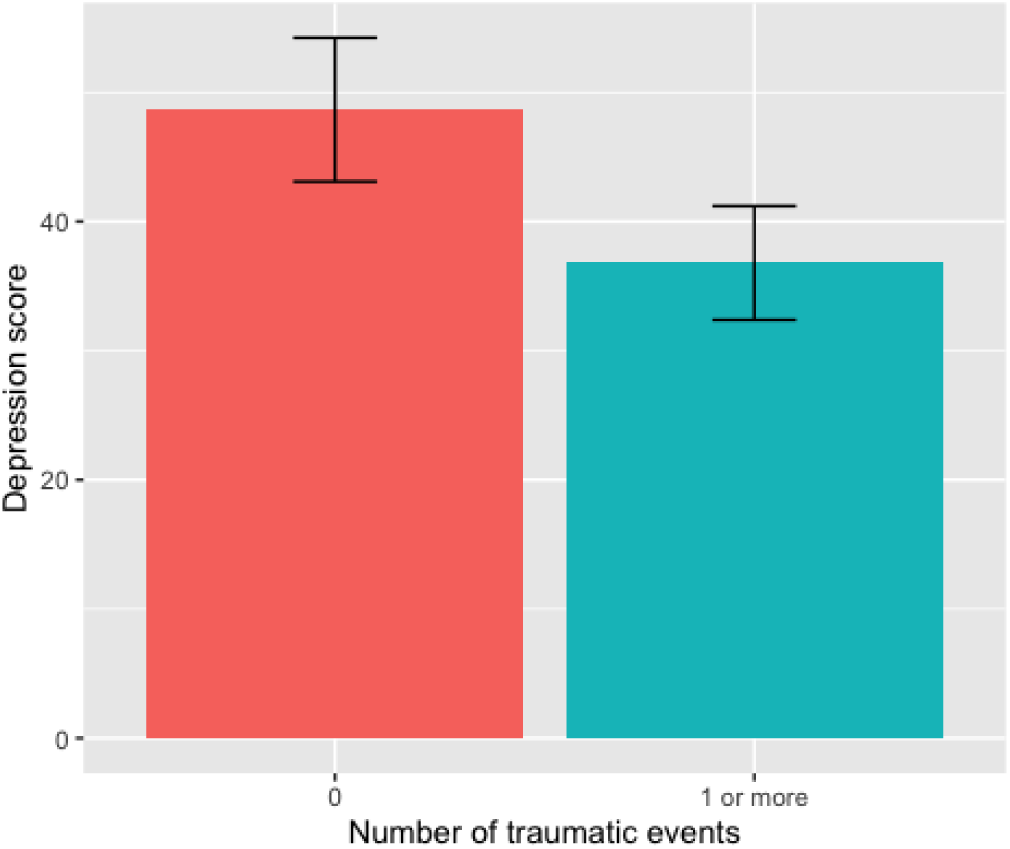
Depression symptoms score (WHO-5) by number of traumatic events

### Depression and economic outcomes

Around one in three Palestinians consider themselves to be poor in 2022. Seventeen percent describe themselves as poor and nine percent as very poor. About half of the participants are currently employed in the labor market, while 20 percent are looking for a job. Among those in employment, the average number of hours worked in the week prior to the interview was 38·9 (SE=0·40) and the average number of months worked in the last year was 9·8 (SE=0·04). The average Food Consumption Score was 74·4 (SE=0·86), which is well above the threshold of 42 for an acceptable FCS. However, six percent of Palestinians in 2022 have a poor or borderline diet. As shown in Figure 5a, (perceived) poverty and depression are strongly linked. For Palestinians who consider themselves as relatively wealthy the mean level is above the threshold of 50. Yet, for all other groups (self-identified as “in the middle”, “poor” or “very poor”), the mean depression level is below the threshold and increases steadily with the level of poverty. Similarly, depression symptoms are significantly worse for those with poor or borderline food security compared to those with acceptable food security (Fig. 5b). Depression is also strongly linked with employment outcomes, at both the extensive and intensive margins. Depression levels are significantly worse among the unemployed than the employed (Fig. 5c). Among the employed, for individuals that engage in greater amounts of work, depression symptoms tend to be less prevalent (Fig. 5d).

**Fig. 5.**
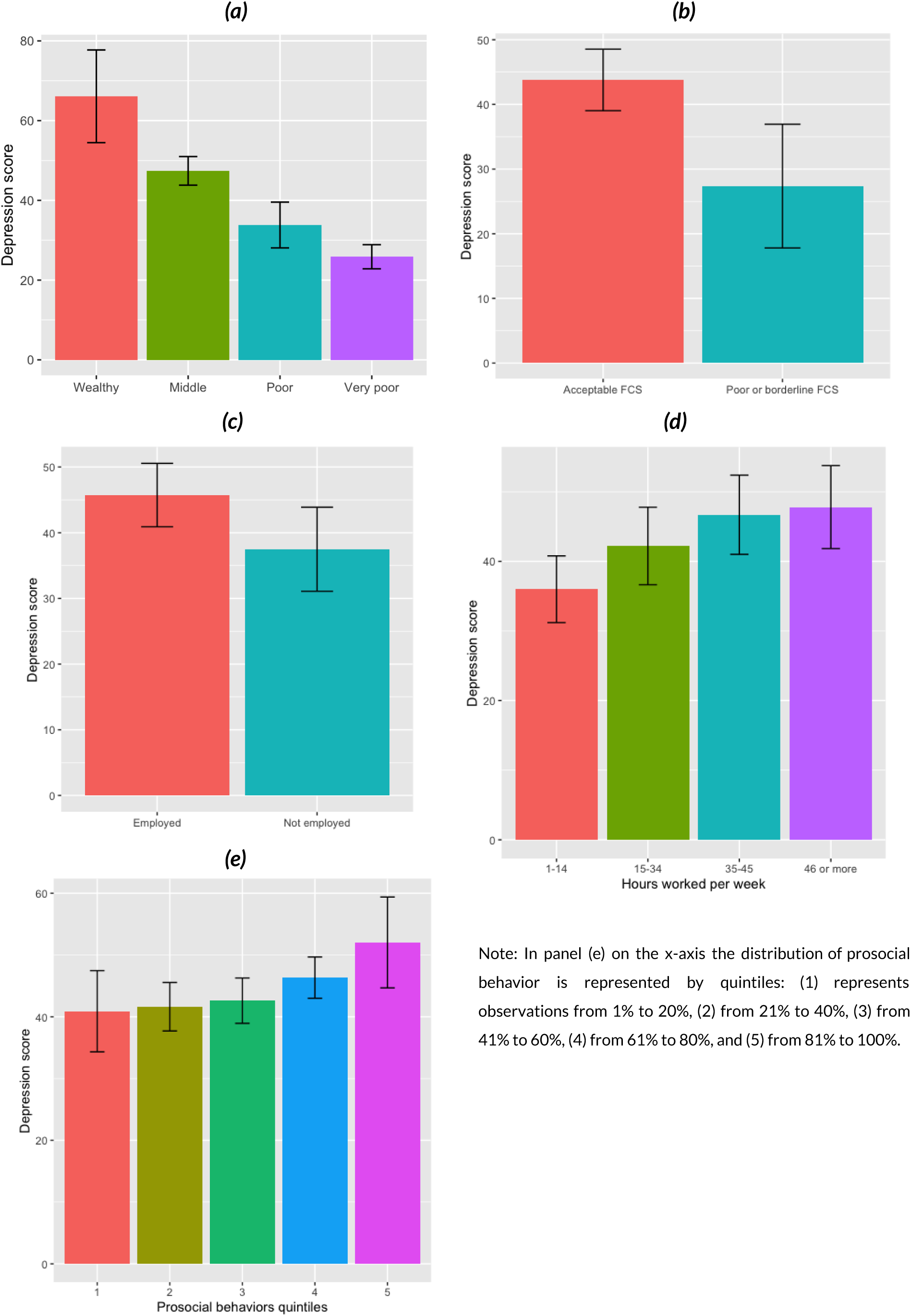
Depression symptoms score by perceived poverty (a), Food Consumption Score (b), employment status (c), hours worked per week (d), and prosocial behaviors quintiles (d)

### Depression and prosocial behavior

We observe that the WHO-5 score increases, indicating a decrease in depressive symptoms, with the number of reported prosocial behaviors in the last six months (Fig. 5e). Particularly for those in the fourth and fifth quintiles, the increase is statistically significant, suggesting that prosociality and healthy participation in social life are associated with lower levels of depressive symptoms.

### Costs of depression

The associated burden of depression is enormous. We estimate that in 2022, depression alone resulted in a depletion of 244,185 Years Lived with Disability (YLD) in the West Bank and 488,370 YLD in Gaza. When weighted on the 2022 per capita income of residents in the West Bank (US$5,300) and Gaza (US$1,440) respectively (World Bank, 2023), this prevalence translates into an approximate economic cost of USD 918 million in the West Bank and USD 703 million in Gaza. On a national scale, the YLD in 2022 tallies up to 732,555 with a corresponding economic cost of USD 1·6 billion. This amount equates to 8·9 percent of Palestine’s GDP for that year. In addition, depression and other mental health issues are often comorbid, as discussed in Appendix A8, suggesting that the above estimates represent the lower bound of costs of depression.

## Discussion

Drawing on our unique data, we demonstrate that in 2022 depression was highly prevalent across Palestine. Our key result is obtained by geospatially linking conflict event data with survey data: we show that the poor mental health of Palestinians is closely linked to conflict exposure. Similarly, self-reported traumatisation and socio-economic deprivation are also linked to depressive symptoms. In a novel calculation for Palestine, we also show that the associated economic losses were enormous, representing about 732,555 Years Lost in Disability and 8·9 percent of Palestine’s GDP in 2022.

Our findings suggest that the most recent increase in violent events since October 7, 2023 may have led to a higher prevalence of depression in Palestine still. The psychological burden of the conflict may exacerbate the negative effect of conflict on the economy, contributing to a likely vicious spiral of poor mental health, high unemployment, and poverty.

Our results reaffirm several previously established observations regarding lives and livelihoods in conflict situations, notably the disproportionate impact of violence on women^27,28^ or the dose-response relationship between conflict exposure and mental health.^29^ Our findings also confirm a large body of evidence documenting gender gaps in depression across contexts.^1^

Our analyses also challenge some existing findings about mental health, for example by documenting that refugees’ mental health levels may not be necessarily worse than that of non-refugees.^29^ In our case, the area of residence (West Bank versus Gaza) is a much stronger predictor of mental health, emphasizing that in active conflicts local conditions play a key role in shaping mental health outcomes. Furthermore, we document a much higher economic burden stemming from depression in Palestine in 2022 than what had been suggested for that of all mental disorders in the MENA region.^30^

Our study also challenges existing methods for capturing conflict exposure and its consequences. Contrasting widely-used self-reported trauma exposure with proximity to conflict events, we find that both trauma and event-based measures suggest negative links with mental health. Yet, the results emphasize that conflict-event based measures may be an important complement to self-reported trauma exposure measures to better grasp the complex challenges conflict situations entail, which should be studied in more depth by future research.

Our results on associations of depression with socio-economic outcomes provide novel and highly policy-relevant findings for conflict settings. Across the full range of analyzed economic and social indicators, depression in times of conflict is associated with worse outcomes, including poverty, employment, and prosociality. These findings underline the serious economic and stability risks conflicts can generate and the important role psychological channels may play in human development. Therefore, there is an urgent need to design, analyze, and scale up investments in mental health research and provision in conflict settings. Our results suggest that for a given level of violence exposure, mental health may be improved through interventions, thus also helping individuals improve their livelihoods. However, livelihood support in isolation, if provided for people living with severe mental health burden, may not be effective on its own; integrated livelihood and mental health interventions in conflict-affected settings may prove to be more effective.

Our study has some limitations. First, while the WHO-5 serves as a widely employed screening tool in numerous international studies and has demonstrated its validity in assessing depression,^22^ the prevalence rate identified within our study should be approached with the customary caution that accompanies the utilization of screening instruments. Second, the cross-sectional character of this study does not allow causal interpretations of our bivariate estimates or analyses of temporal dynamics. We leave such analyses to possible further waves of the Palestinians’ Psychological Conditions Survey (PPCS), which would provide a novel, nationally representative panel dataset of mental health in an acute conflict setting.

## Data Availability

All data produced in the present study are available upon reasonable request to the authors

## Online appendices

### A1 Literature on depression and conflict

The prevalence of depression in conflict-affected areas is widely reported. Charlson et al.^1^ reviewed 83 studies of conflict-affected populations and found a prevalence of major depression of 7·6 percent, which is more than twice the mean prevalence reported by the GBD Study (3·5 percent for major depressive disorder, MDD). In a recent meta-analysis Lim et al.^2^ highlight an even higher prevalence of 33·3 percent for depression among war-affected civilians, similar to the estimated 27 percent prevalence among civilian war survivors found in a meta-analysis by Morina et al.^3^ Hoppen and Morina^4^ note that more than 300 million adult war survivors suffer from PTSD and/or major depression. Moradinazar et al.^5^ studied the burden of depressive disorders in the MENA region using the GBD database and found the highest prevalence, incidence, and disability-adjusted life years in Palestine. A similar study has been conducted by Safiri et al.,^6^ reiterating the finding that Palestine accounts for the largest share of the burden of major depressive disorders in the Middle East and Northern Africa region. An existing study by Veronese et al.^7^ (2017) on Palestinian professional helpers from the Gaza and West Bank (N=201) found a mean depression score of 41 (SD=22·84). In a small but nationally representative sample of Palestinian adults, Canetti et al.^8^ showed that screening for depression is positively associated with exposure to political violence and with greater loss of interpersonal and intrapersonal resources or the loss of a loved one. Focusing only on Gaza, several scholars examined the effect of exposure to conflict and violence on physical health^9,10^ and revealed how people residing in areas where there are more instances of violence are more likely to have physical disabilities and chronic illnesses, in particular mental health issues, compared to others that have not experienced violence. These findings suggest that exposure to conflict and violence increases the risk of developing or maintaining depression. Concurrently, prosocial behaviors, such as acts of kindness, empathy, and cooperation towards others, have been consistently linked to various mental health benefits, including improved well-being, reduced stress, and increased social support.^11^

Depression is globally an undertreated condition, with people in low-income countries generally receiving less treatment than those in high– and middle-income countries^12^ (Thornicroft et al., 2017), resulting in a huge treatment gap. This becomes also apparent when looking at the mental health workforce of these countries. In Palestine, there were 0·9 psychiatrists and 1·0 psychologists per 100,000 inhabitants in 2007.^13^ In high income countries, the mental health workforce is significantly higher. In 2016, for example, in the United States it was 10·5 and 29·9, respectively, and 13·2 and 49·6, respectively, in Germany^14^ (WHO, 2023). This further underlines the gravity of our findings. Nevertheless, further research is needed to better understand these relationships.

### A2 Sampling design and estimation methodology

The Palestinians’ Psychological Conditions Survey (PPCS) is a face-to-face panel survey conducted by the Palestinian Central Bureau of Statistics (PCBS) in March and April 2022. The PPCS sample was designed as a randomly selected subsample of the respondent sample of the first round of the COVID-19 Rapid Assessment Phone Survey (RAPS 1), which was fielded by PCBS between June and August of 2020. The RAPS survey, in turn, was a continuation of the latest panel wave of the Socio-Economic and Food Security Survey (SEFSEC 2018). Consequently, the sample of the 2022 PPCS survey can be regarded as a new (partial) wave of the SEFSEC panel initiated in 2013. Longitudinal information spanning such an extended time is extremely valuable for impact evaluation and causal inference. At the same time, panel surveys can be considered fully representative of their target population only at round one, whereas their cross-sectional representativeness may decay as the panel ages, because of attrition, life-cycle dynamics of the panel members, and structural changes in the target population. Specific weighting techniques have been applied to counteract those effects in the case of the PPCS sample, thereby improving its ability to provide a reliable representation of 2022 Palestine. In this respect, it is important to note that both panel ancestors in the lineage of the PPCS sample (namely SEFSEC and RAPS) were considered by PCBS to be representative at national, governorate, and rural/urban/camps levels at the time of the first wave.

The planned sample size for PPCS was set to 7,057 households, with the 8,709 respondent households of RAPS 1 serving as the sampling frame. The selection of PPCS households followed a one-stage cluster sampling design. Specifically, 641 Enumeration Areas (EA) were randomly selected from the 1,824 EAs of RAPS 1 with Probability Proportional to Size (PPS). All the RAPS 1 households contained in the selected EAs were included in the PPCS sample. To accommodate logistical needs of data collection, variable “number of RAPS 1 respondent households per EA” was used as measure of size (MOS) for the PPS algorithm. This made the inclusion of EAs containing fewer RAPS 1 households less likely, thus minimizing the number of EAs to be visited and hence alleviating organizational challenges and data collection costs. Nonetheless, every household in the sampling frame received a known and strictly positive probability of being included in the PPCS subsample, which fully preserved the probability sampling nature of the PPCS survey. This deserves to be emphasized, because probability sampling is the essential ex-ante guarantee for any survey to generate unbiased estimates of the target population.

The individual questionnaire of the PPCS survey, through which mental health information was collected, was administered to one selected adult member (aged 18 years or above) of each respondent household. For each respondent PPCS household, interviewers attempted to identify and re-interview the same adult individual who responded to the individual module of SEFSEC 2018. Only if the attempt was unsuccessful, the interviewer used a Kish grid (which accompanied the questionnaire) to randomly select, with equal probability, one adult from among all adult members of the household. The re-interview attempt was successful in 91 percent of the cases. Of 7,057 planned households, 917 did not respond, yielding an overall household-level response rate of 87 percent. Of those who did not respond, in 84 percent of cases the household was not found or had moved abroad, in 12 percent of cases the household refused to be interviewed again, and the remaining households only partially completed the interview. Out of the 6,140 respondent households, 5,877 adult individuals completed the mental health module of the questionnaire. Therefore, the individual-level response rate, conditional on the respondent household sample, was 96 percent.

Since nonresponse results in risk of bias and loss of precision in estimation, PPCS weights were carefully adjusted for both household-level and individual-level nonresponse. Given the origin of the PPCS sample — that is, its provenance from the RAPS and SEFSEC panels — rich information was available on both respondents and non-respondents, which made it possible to calculate nonresponse adjustments by adopting a robust response-propensity modeling approach. After the adjustment for nonresponse, the household-level and individual-level weights were also calibrated to obtain an additional layer of protection against bias and enhance estimation efficiency. This calibration adjustment allows PPCS to exactly match in estimation 163 known population totals about the Palestinian population in 2022, thereby solidifying its cross-sectional representativeness. The known population totals employed as calibration benchmarks are (i) counts of Palestinian households by governorate and locality type (43 totals), (ii) counts of Palestinian persons by region, five-year age class, and sex (68 totals), and (iii) counts of Palestinian *adult* persons by region, five-year age class, and sex (52 totals). Finally, the obtained calibration weights were trimmed to get rid of unduly large weights that might have led to unstable estimates and inflated standard errors. As a further bias prevention safeguard, the trimming adjustment was performed consistently, that is simultaneously preserving all the calibration constraints. Overall, the fundamental objectives that informed the calculation of PPCS weights were mitigation of bias risks and improvement of estimation efficiency.

The obtained household-level and individual-level weights have been publicly disseminated together with the validated PPCS microdata (available at the link https://microdata.worldbank.org/index.php/catalog/5807). Unless otherwise stated, the point estimates reported throughout this paper are based on those weights, and the related standard errors fully account for the complex sampling design of the PPCS survey.

### A3 Additional information on survey measures

#### Depression

The WHO-5 comprises five positively stated statements, which respondents rate in reference to their previous two weeks using a six-point Likert scale ranging from five “All of the time” to zero “At no time”. We calculate the final score as the sum over the five items (range: 0-25) multiplied by 4 (range: 0-100). A score below or equal to 50 is classified as a “screening diagnosis” for depression.^15^

#### Traumatic events

To capture exposure to conflict and trauma in the survey, we adapted the Brief Trauma Questionnaire (BTQ) to the Palestinian context.^16,17^ Respondents were asked if they had experienced any of ten potentially traumatic events that could be answered with “yes” or “no”. Questions included for example whether individuals have been exposed to or witnessed war-related casualties, or whether they have witnessed a situation in which someone was seriously injured or killed (see Appendix A6 for the full list).

#### Proximity to conflict events

To measure proximity to conflict events, we spatiotemporally match PPCS data with geo-tagged and time-stamped conflict event data from the Armed Conflict Location & Event Data Project (ACLED).^18^ ACLED tracks a range of violent and non-violent actions by political actors, including governments, rebels, militias, identity groups, political parties, external actors, rioters, protesters, and civilians. We then count the number of conflict events that occurred within two, five, ten and fifty kilometers of a household’s location one and five years before the interview. The relatively small radius and time interval are deliberately chosen to ensure a nuanced reflection of the complex and localized nature of conflict exposure in Palestine. This approach is particularly important in contexts where conflict events tend to concentrate on precise areas, such as the Jenin refugee camp and the old city of Hebron. The advantage of a small radius becomes also evident in densely populated areas, such as Gaza, where geographical constraints require a more fine-grained analysis.

#### Classification of ACLED events

The Armed Conflict Location & Event Data Project (ACLED), uses a global methodology to track a range of violent and non-violent actions by political agents, including governments, rebels, militias, identity groups, political parties, external actors, rioters, protesters, and civilians (ACLED, 2019).^19^ This study uses the ACLED codebook to categorize the violent events that were registered in the West Bank and Gaza one and five years before data collection. Violent events include battles, explosions/remote violence, and violence against civilians. Table A.1 describes the full set of sub-event considered and Table A.2 reports the frequencies of violent events for each governorate in the two time periods examined.

**Table A.1:** ACLED classification of violent events.

| Classification | Event type | Sub-event type |
| --- | --- | --- |
| Violent events | Battles | Armed clash |
|  |  | Government regains territory |
|  |  | Non-state actor overtakes territory |
|  | Explosions/ Remote violence | Chemical weapons |
|  |  | Air/drone strike |
|  |  | Suicide bomb |
|  |  | Shelling/artillery/landmine/IED |
|  |  | Remote explosive |
|  |  | Grenade |
|  | Violence against civilians | Sexual violence |
|  |  | Attack |
|  |  | Abduction/ forced disappearance |

**Table A.2:** Frequencies of violent events (ACLED) in Palestine one and five years before the survey.

| Governorate | One year before the survey | Five years before the survey |
| --- | --- | --- |
|  | Battles, Explosions, and Violence vs Civilians | Battles, Explosions, and Violence vs Civilians |
| Bethlehem | 23 | 119 |
| Deir El Balah | 23 | 257 |
| Gaza City | 85 | 440 |
| Hebron | 78 | 277 |
| Jenin | 56 | 149 |
| Jericho | 7 | 22 |
| Jerusalem | 142 | 344 |
| Khan Yunis | 36 | 295 |
| Nablus | 58 | 178 |
| North Gaza | 62 | 435 |
| Qalqilya | 14 | 79 |
| Rafah | 19 | 181 |
| Ramallah and Al Bireh | 48 | 197 |
| Salfit | 15 | 61 |
| Tubas | 10 | 27 |
| Tulkarm | 16 | 82 |
| Total | 692 | 3,143 |

#### Years Lived with Disability

Following the approach proposed by Lipton et al.^20^ and adopted by OECD we used the average disability weight for depressive disorder of 0·219 (for individuals with WHO-5 score ≤50). The utilization of the average disability weight offers the benefit of not requiring additional assumptions about the WHO-5 cutoffs, as the WHO-5 serves as a screening tool and isn’t intended to differentiate levels of depressive disorder severity. The economic loss attributable to depression is calculated by multiplying the YLD value by the 2022 GDP per capita of residents in the West Bank (US$5,300) and Gaza (US$1,440) respectively.^21^ We computed the economic loss separately for the two areas to account for the structurally different economic conditions. Doing so, our estimates are conservative because they partially rule out the second order effect of poor mental health in economic performance (i.e. the fact that in Gaza part of the poor economic performance is due to the higher prevalence of depression).

#### Food Consumption Score

FCS reflects the current state of food security (last seven days) and is ideal for tracking changes over time. Subjects are asked about the frequency of consumption in days over a seven-day recall period. Food items are classified into standard food groups and have a maximum value of seven days per week. The FCS is calculated by multiplying the frequency of consumption of each food group by an assigned weight based on its nutrient content, resulting in an index ranging from zero to 112. To create the profiles, we used daily oil and sugar intake thresholds (around seven days per week) that reflect the most common diet among respondents. Accordingly, three profiles are identified: poor food consumption for values less than 28, borderline food consumption for values between 28 and 42, and acceptable food consumption for values greater than 42. Given the low number of respondents who fall into the poor food consumption category, for the binary FCS variable the two categories “poor” and “borderline” were combined into the category: “poor or borderline”.

#### Prosocial behavior

Using a 10-item scale, participant were asked whether or not they have taken the following actions in the previous half year: “Made a financial contribution to a charity, zakat or philanthropic organization”; “volunteered your time at a charity or philanthropic organization”; “Allowed someone to go ahead of you in a queue”; “Loaned someone money”; “Loaned someone an item”; “Carried someone’s belongings”; “Given directions”; “Helped someone find a job”; and “Given money to a beggar or purchased something you did not need from a street seller in order to help them (bought food for someone)”.

### A4 Socio-demographic statistics

Table A.3 presents an overview of the unweighted sample characteristics. The respondents’ average age is 44 years (SD=16·15, ranging from 18 to 99 years). Of the respondents 50 percent are men, around twenty percent are unemployed, and among those working they worked on average 38·9 (SD=0·4) hours in the week before the interview. 40 percent have refugee status. About one in three Palestinians consider themselves to be economically poor, with seventeen percent describing themselves as poor and nine percent as very poor.

**Table A.3:**
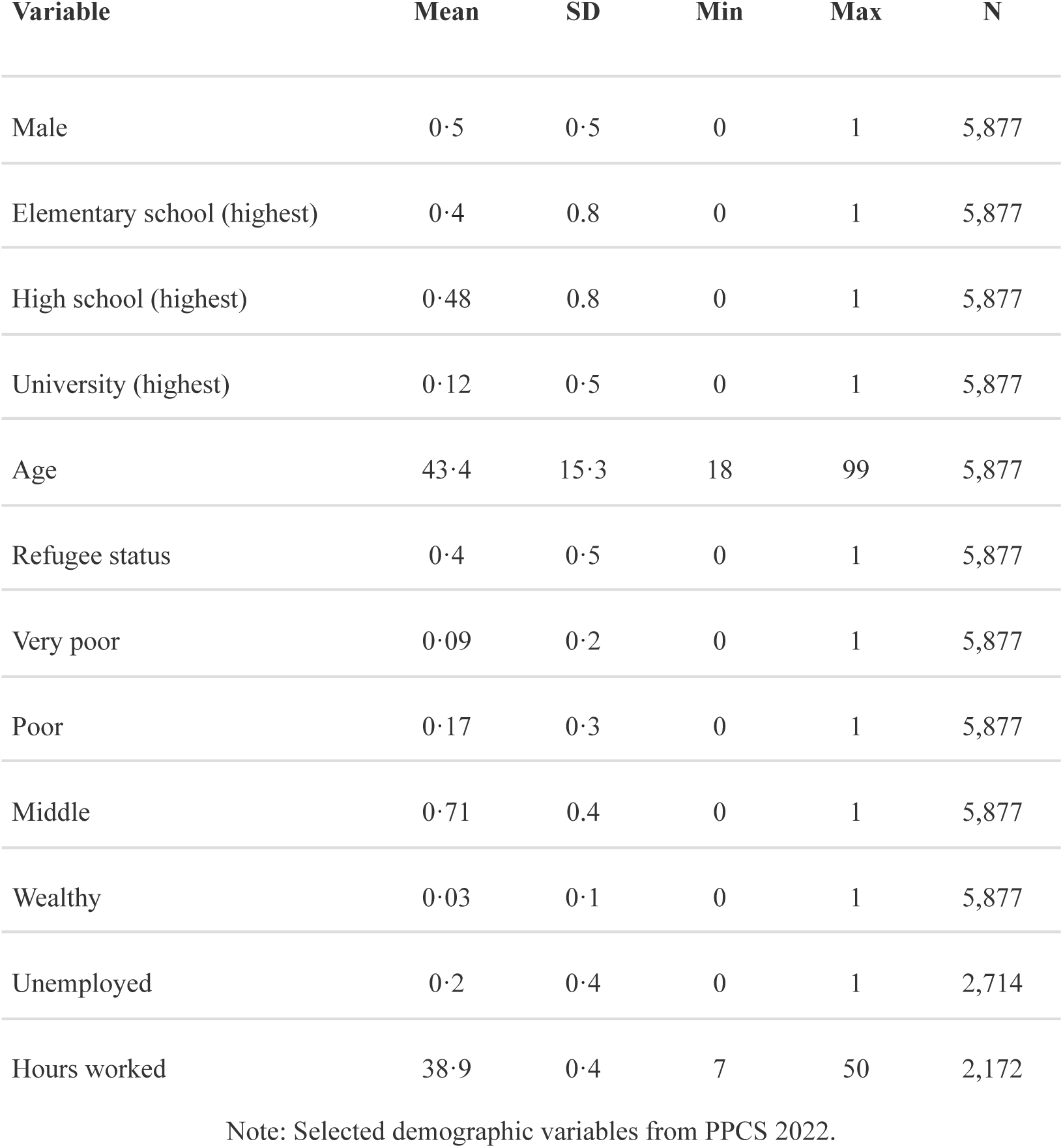
Sample characteristics.

| Variable | Mean | SD | Min | Max | N |
| --- | --- | --- | --- | --- | --- |
| Male | 0.5 | 0.5 | 0 | 1 | 5,877 |
| Elementary school (highest) | 0.4 | 0.8 | 0 | 1 | 5,877 |
| High school (highest) | 0.48 | 0.8 | 0 | 1 | 5,877 |
| University (highest) | 0.12 | 0.5 | 0 | 1 | 5,877 |
| Age | 43.4 | 15.3 | 18 | 99 | 5,877 |
| Refugee status | 0.4 | 0.5 | 0 | 1 | 5,877 |
| Very poor | 0.09 | 0.2 | 0 | 1 | 5,877 |
| Poor | 0.17 | 0.3 | 0 | 1 | 5,877 |
| Middle | 0.71 | 0.4 | 0 | 1 | 5,877 |
| Wealthy | 0.03 | 0.1 | 0 | 1 | 5,877 |
| Unemployed | 0.2 | 0.4 | 0 | 1 | 2,714 |
| Hours worked | 38.9 | 0.4 | 7 | 50 | 2,172 |

Note: Selected demographic variables from PPCS 2022.

### A5 Exposure to conflict events

**Table A.4:** Conflict event exposure.

| Variable | Mean | SD | Min | Max | N |
| --- | --- | --- | --- | --- | --- |
| Radius: 2 km; period: one year | 7.92 | 11.98 | 0 | 65 | 5,877 |
| Radius: 5 km; period: one year | 11.44 | 15.15 | 0 | 79 | 5,877 |
| Radius: 10 km; period: one year | 30.76 | 34.08 | 0 | 119 | 5,877 |
| Radius: 50 km; period: one year | 136.14 | 55.57 | 15 | 225 | 5,877 |
| Radius: 2 km; period: five years | 44.54 | 68.81 | 0 | 352 | 5,877 |
| Radius: 5 km; period: five years | 64.83 | 86.71 | 0 | 419 | 5,877 |
| Radius: 10 km; period: five years | 171.67 | 194.98 | 0 | 716 | 5,877 |
| Radius: 50 km; period: five years | 716.25 | 422.67 | 44 | 1,612 | 5,877 |

Note: ACLED violence events matched with data from PPCS 2022. Periods refer to years before the date of the survey.

### A6 Exposure to traumatic events

**Table A.5:** Personal traumatic events in Palestine (lifetime)

| Traumatic events | count<br>(yes) | percent<br>(yes) |
| --- | --- | --- |
| Have you ever been exposed to or witnessed war-related casualties (for example as a medic or on graves registration duty)? | 1708 | 29·1 |
| Have you ever been in a serious car accident, or a serious accident at work or somewhere else? | 745 | 12·7 |
| Have you ever been in a major natural or technological disaster, such as a fire, tornado, hurricane, flood, earthquake, or chemical spill? | 218 | 3·7 |
| Have you ever had a life-threatening illness such as cancer, a heart attack, leukemia, AIDS, multiple sclerosis, etc.? | 311 | 5·3 |
| Before age 18, were you ever physically punished or beaten by a parent, caretaker, or teacher so that: you were very frightened; or you thought you would be injured; or you received bruises, cuts, welts, lumps or other injuries? | 769 | 13·1 |
| Not including any punishments or beatings you already reported In Question 5, have you ever been attacked, beaten, or mugged by anyone, including friends, family members, or strangers? | 306 | 5·2 |
| Has anyone ever made or pressured you into having some type of unwanted sexual contact?<br>Note: By sexual contact we mean any contact between someone else and your private parts or between you and some else's private parts | 34 | 0·6 |
| Have you ever been in any other situation in which you were seriously injured, or have you ever been in any other situation in which you feared you might be seriously injured or killed? | 296 | 5·0 |
| Has a close family member or friend died violently, for example, in a serious car crash, mugging, or attack? | 1005 | 17·1 |
| Have you ever witnessed a situation in which someone was seriously injured or killed, or have you ever witnessed a situation in which you feared someone would be seriously injured or killed? | 569 | 9·7 |

### A7 Exposure to conflict events and traumatic events

**Table A.7:** Correlation of exposure to conflict events with exposure to traumatic events.

| Variables | 1 year | 5 years | N |
| --- | --- | --- | --- |
| Radius: 2 km | ·088 | ·118 | 5,877 |
| Radius: 5 km | ·101 | ·127 | 5,877 |
| Radius: 10 km | ·132 | ·167 | 5,877 |
| Radius: 50 km | ·045 | ·181 | 5,877 |

Note: Traumatic events as reported in the Brief Trauma Questionnaire (BTQ) as part of the PPCS 2022. ACLED violence events matched with data from PPCS 2022. Periods refer to years before the date of the survey.

### A8 Depression and other mental health issues

Depression, anxiety disorders, and post-traumatic stress disorder (PTSD) are often comorbid mental health issues with complex interdependencies. Studies have shown that individuals with depression are at an increased risk of developing anxiety disorders and PTSD, and vice versa.^22^ Common environmental risk factors, such as exposure to traumatic events or chronic stress, can contribute to the development of depression, anxiety disorders, and PTSD and the presence of one disorder may also exacerbate symptoms of the other conditions, leading to a cycle of increased psychological distress.^23^ Moreover, overlapping symptomatology, such as sleep disturbances, intrusive thoughts, and emotional dysregulation, further highlights the interdependencies between these mental health issues.

**Fig. A.1:**
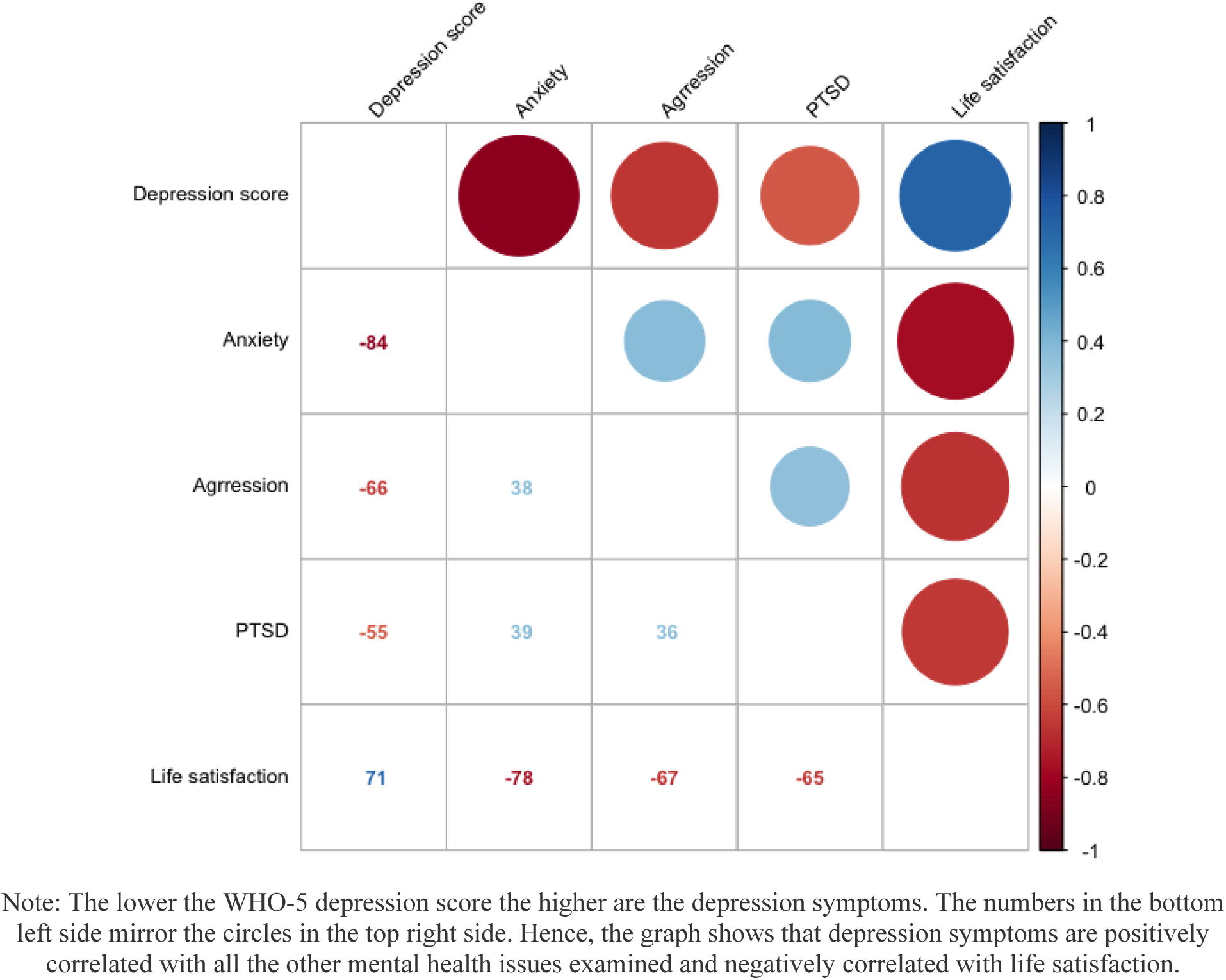
Correlation matrix of mental health issues

## Footnotes

1 For the 50km radius, we only consider correlations between conflict events in Gaza and the West Bank with depression scores in Gaza and West Bank, respectively. That is, we exclude the few cases where conflict events in Gaza may have correlated with depression scores in the West Bank and vice versa.

2 We also estimated the association between violence events within a 1km radius and depression but in that case the number of individuals exposed to no conflict events is fairly large (53 percent). As a result, estimation of associations with depression is more imprecise (but qualitatively similar to the results for a 2km radius).

## Notes

### Competing Interest Statement

The authors have declared no competing interest.

### Author Declarations

The study protocol was approved by An-Najah National University Institutional Review Board (IRB) Committee with all research performed in accordance with the relevant guidelines and regulations.

### Summary of Updates

abstract now displayed in the correct format

